# The interaction between coping styles with relationship breakdowns, loss, and conflict and the frequency of self-harm thoughts and behaviours: a longitudinal analysis of 21,581 UK adults

**DOI:** 10.1101/2022.06.21.22276696

**Authors:** Elise Paul, Daisy Fancourt

## Abstract

**Background:** Relationship breakdowns or conflict are frequent precipitants for self-harm thoughts and behaviours, but the majority who experience these stressful life events do not think about or engage in self-harm. Understanding factors that attenuate or exacerbate this risk is therefore needed. The aim of this paper is to investigate whether relationship breakdowns, loss, and conflict lead to more frequent self-harm thoughts and behaviours. We also examine whether coping styles and neuroticism, posited by diathesis-stress models of suicide risk to interact with these events, attenuate or exacerbate the risk for self-harm thoughts and behaviours from these events.

**Methods:** This study utilised data collected during the COVID-19 pandemic, which acted as a natural experiment by leading to a greater prevalence of relationship breakdowns than usual. Data from 21,581 adults who participated in the UCL COVID-19 Social Study between 28 February 2021 and 4 April 2022 were utilised. Poisson regression models which controlled for socio-demographics and a diagnosed mental health condition were used to examine the impact of four predictor variables (separation or divorce, family problem, an ‘other’ relationship breakdown [e.g., friend or colleague], and the death of a close relative or friend) with the number of times self-harm thoughts and behaviours were reported over the study period. Interactions between these events and coping styles (problem-focused, emotion-focused, socially supported, and avoidant coping) were also examined.

**Results:** Variables representing relationship breakdowns, loss, and conflict were associated with an increased frequency of self-harm thoughts (incidence rate ratio [RRR] range: 1.04 to 1.77) and behaviours (RRR range: 1.48 to 1.96). The use of more avoidant coping strategies (e.g., substance use, denial) increased the risk for both outcomes but unexpectedly attenuated associations between predictor variables and self-harm thoughts. Socially supported coping increased the likelihood of both outcomes, but not in sensitivity analyses which excluding ‘venting’ from the scale. Problem-focused coping strategies (e.g., active planning) attenuated the impact of separation or divorce and having had a family problem on the frequency of self-harm behaviours.

**Conclusions:** Findings underscore the importance of interpersonal loss and conflict for the frequency of thinking about and engaging in self-harm and suggest that the magnitude of these associations may depend on different coping styles.

## Background

Self-harm thoughts and behaviours are significant public health problems and significant risk factors for future death by suicide,^1–3^ particularly when repeated.^4, 5^ Relationship difficulties with romantic partners and family members are the most frequent stressors precipitating self-harm,^6, 7^ and diathesis-stress models of self-harm thoughts and behaviours emphasise the importance of these life events as proximally occurring risk factors for self-harm.^8, 9^ However, the vast majority of people who experience a relationship breakdown do not think of death or harming themselves.

According to diathesis-stress models of suicide and self-harm risk, stable and enduring factors such as maladaptive ways of coping with stress form a long-term vulnerability to thinking about and engaging in self-harm (the diathesis).^8, 10^ Acute risk for self-harm is then greatest when stressful life events such as relationship breakdowns and conflict act on the underlying diathesis to exacerbate their impact.^8, 9^ Different coping styles, or the cognitive and behavioural strategies individuals use to deal with stress and adversities,^11^ have shown differential associations with self-harm risk. Coping strategies which are characterised by avoiding a stressful situation or difficult experience (e.g., using denial, distraction or using drugs or alcohol), so-called ‘avoidance strategies’, have been shown to increase the likelihood of self-harm thoughts and behaviours and other mental health problems,^12, 13^ whilst problem oriented coping (e.g., taking steps to actively make the situation better), emotion-focused coping (e.g., positive reframing), and emotional support from others (socially supported coping) decrease their likelihood.^14, 15^

However, despite evidence for these direct associations with self-harm thoughts and behaviours, hardly any research has examined interactions between these long-term factors and stressful life events such as relationship dissolution and conflict occurring in closer proximity to self-harm thoughts and behaviours. The COVID-19 pandemic provides a natural experiment through which to explore this topic. It placed additional strains on interpersonal relationships that may have resulted in more frequent relationship breakdowns and conflicts compared to before the pandemic. For example, there were reported increases across the pandemic in relationship dissolution and conflict.^16, 17^ But research testing whether these stressors interact with longer-standing influences to increase or decrease risk for self-harm thoughts and behaviours is lacking. Further, evidence for how these risk factors influence the frequency of self-harm thoughts and behaviours over time is needed, as most studies of self-harm thoughts and behaviours have been insufficiently powered. This is especially important because risk for future death by suicide is especially strong when self-harm thoughts and behaviours are more frequent.^5^

This study directly addresses these needs by examining whether relationship breakdowns, loss and conflict were associated with increasing frequency of self-harm thoughts and behaviours over the course of the second year of the pandemic in the UK. We chose the second year of the pandemic as it presented a period after the initial psychological shock had passed and during which time there were fewer social restrictions than in Year 1, thereby presenting more ordinary patterns of social behaviours. We then explored whether these associations between relationship factors and self-harm thoughts and behaviours were attenuated or exacerbated by four different coping styles: emotion focused, problem focused, socially supported, and avoidant coping. The results have the potential to deepen our understanding of the circumstances in which relationship breakdowns, loss, and conflict lead to self-harm thoughts and behaviours and to make clinical assessments more meaningful.

## Methods

### Study design and participants

Data were drawn from the COVID-19 Social Study; a large panel study of the psychological and social experiences of over 75,000 adults (aged 18+) in the UK during the COVID-19 pandemic. The study commenced on 21 March 2020 and involves online data collection from participants for the duration of the COVID-19 pandemic in the UK. Data were initially collected weekly (from study commencement through August 2020), then monthly thereafter. The study is not random and therefore is not representative of the UK population. But it does contain a well-stratified sample that was recruited using three primary approaches. First, convenience sampling was used, including promoting the study through existing networks and mailing lists (including large databases of adults who had previously consented to be involved in health research across the UK), print and digital media coverage, and social media. Second, more targeted recruitment was undertaken focusing on groups who were anticipated to be less likely to take part in the research via our first strategy, including (i) individuals from a low-income background, (ii) individuals with no or few educational qualifications, and (iii) individuals who were unemployed. Third, the study was promoted via partnerships with third sector organisations to vulnerable groups, including adults with pre-existing mental health conditions, older adults, carers, and people experiencing domestic violence or abuse. The study was approved by the UCL Research Ethics Committee [12467/005] and all participants gave informed consent. Participants were not compensated for participation (https://osf.io/jm8ra/).

Our study focused on the second year of the pandemic, from 28 February 2021 to 4 April 2022. We included participants who met the following criteria. First, participants were included if they had participated in the survey between 21 March and 4 April 2022, when we measured the self-harm thoughts and behaviours outcome and relationship breakdown outcome variables (N = 29,337), and had to have non-missing data on these outcomes (N = 28,583). Second, participants had to have non-missing data on the coping styles module, which was administered in May 2021 (N = 23,609). Third, participants were included if they had non-missing data on variables collected at baseline: those required to calculate statistical weights (gender, age, ethnicity, country, and education), and whether they had been diagnosed with a mental health condition (N = 22,769). Fourth, because we were interested in looking at the frequency of self-harm thoughts and behaviours over the study period, we included only participants who also had data on these two variables for at least three time points over the course of the year prior to the date of their March 2022 survey (+/- 28 days) (N = 21,581). This criterion was what generated the specific study period dates of 28 February 2021 to 4 April 2022. Participants included in the study provided an average of 10.17 assessments (SD = 2.68). See Supplemental Table S1 for a comparison of excluded and included participants on study variables.

### Measures

#### Outcome variables

Two outcome variables were used to measure the frequency of self-harm thoughts and self-harm behaviours in the past week, assessed with one question each at the time of the March 2022 survey. Self-harm thoughts were measured with an item from the Patient Health Questionnaire (PHQ-9);^18^ an instrument often used as a screening tool for depression in primary care practice: “Over the last week, how often have you been bothered by thoughts that you would be better off dead or hurting yourself in some way?”. Second, self-harm behaviours were measured with a study-developed item designed to be similar in wording: “Over the last week, how often have you been bothered by self-harming or deliberately hurting yourself?”. Responses to both items were rated on a four-point scale from “not at all” to “nearly every day”, and dichotomised into none vs any at each assessment point over the study period.

#### Exposure variables

Four predictor variables were adapted from the Life History Interview from the English Longitudinal Study of Ageing^19^. They were administered 21 March to 4 April 2022 about experiences participants may have had in the 12 months prior: i) Divorce, separation, or break-up of personal intimate relationship, ii) Other marital or family problem, iii) Breakdown of another relationship (e.g., with a friend or colleague), and Death of a close relative or friend. Participants rated the extent to which each life event had upset them on a five-point scale from no, it didn’t happen to yes, but it didn’t upset me at all. See Supplemental Table S2 for question and response option wording. A binary variable was created to compare people to whom the event did not happen vs all other four response options. As a sensitivity check, analyses were re-run with binary variables comparing those to whom the breakdown did not happen, or who were not upset too much or not upset at all vs those who were moderately or very much upset by the breakdown.

#### Potential moderators

Four variables were examined as moderators between the relationships between relationship breakdowns and outcomes. Coping styles were measured using the 28-item Brief COPE^20^ scale, which measures the ways in which people typically respond to and cope with stress. The scale was administered 3 to 30 May 2021. When coping data were not available for participants at this date, coping data from the same scale which was administered 7 to 14 May 2020 was used. Most study participants provided coping data from 2021 (90.6%), leaving just 2,026 participants for sensitivity analyses which would exclude people for whom 2020 coping data were used.

A previously-derived four factor model was used for our analyses: i) Problem-focused coping (active coping, planning), ii) Emotion-focused coping (positive reframing, acceptance, humour, religion), iii) avoidant coping (behavioural disengagement, denial, substance use), and iv) socially supported coping (emotional support, instrumental support, and venting).^21^ Because venting has been shown to have a negative impact on mental health,^12, 22^ we conducted sensitivity analyses with the two venting items removed from the socially supported coping scale.

#### Covariates

Socio-demographic factors included were gender (male vs female), age (18-29 vs 30-44 vs 45-59 vs 60+), ethnicity (white vs ethnic minority groups [see Supplemental Table S2 for a full listing of response options]), and education (undergraduate or higher vs A-Levels or vocational training vs up to GCSE). A binary variable constructed from participant responses at baseline to indicate the presence of a clinically diagnosed mental health condition was also included (Supplemental Table S2).

#### Analyses

First, four Poisson regression models were conducted with all four exposure variables entered simultaneously as predictors of self-harm thoughts and behaviours frequency over the past year: without covariates and then with covariates. Second, these Poisson regressions were conducted with interaction terms between the exposure variables and the four moderator variables. In these analyses, covariates were included and linear terms for the moderators were included that were mean-centred. Regression coefficients were exponentiated and reported as incidence rate ratios (IRR). To increase representativeness of the UK general population, data were weighted to the proportions of gender, age, ethnicity, country, and education obtained from the Office for National Statistics.^23^ A multivariate reweighting method was implemented using the Stata user written command ‘ebalance’,^24^ and analyses were conducted using Stata version 17.^25^

## Results

### Descriptive characteristics

Before statistical weights were applied, the sample was disproportionately female, from white ethnic backgrounds, and held at least a university degree or higher, but following weighting the sample aligned well with population proportions (Supplemental Table S3). Just over 1 in 5 (21.1%) of the sample had reported thoughts of self-harm over the study period (mean number of reports: 4.46, SE: 0.09]), whilst 5.6% reported having self-harmed at least once (mean number of reports: 6.01, SE: 0.18]), (Table 1). Most people who had self-harmed at least once had also reported self-harm thoughts (91.7%), but only 1 in 4 who had thought about self-harm had engaged in it (25.0%).

**Table 1.** Sample characteristics for the total sample and by whether self-harm thoughts and behaviours had been reported at least once over the study period, weighted

|  | Total sample<br>N = 21,581<br>Prop. /M (SE) |  | Participants who<br>reported self-harm<br>thoughts at least<br>once<br>N = 4,497<br>Prop. /M (SE) |  | Participants who<br>reported self-harm<br>behaviours at least<br>once<br>N = 1,109<br>Prop. /M (SE) |  |
| --- | --- | --- | --- | --- | --- | --- |
| Self-harm thoughts at least once over the study period | 21.1% |  | -- |  | 91.7% |  |
| Number of times self-harm thoughts were reported over the study period | 0.94 | 0.03 | 4.46 | 0.09 | 6.01 | 0.18 |
| Self-harm behaviours at least once over the study period | 5.6% |  | 25.0% |  | -- |  |
| Number of times self-harm behaviours were reported over the study period | 0.19 | 0.01 | 0.87 | 0.06 | 3.43 | 0.16 |
| Female (ref male) | 50.8% |  | 53.0% |  | 55.9% |  |
| Age (ref 60+) |  |  |  |  |  |  |
| 45-59 | 29.1% |  | 33.4% |  | 37.1% |  |
| 30-44 | 14.4% |  | 20.8% |  | 19.3% |  |
| 18-29 | 5.8% |  | 10.4% |  | 14.9% |  |
| Ethnic minority groups (ref White) | 6.5% |  | 7.8% |  | 7.6% |  |
| Education (ref degree or higher) |  |  |  |  |  |  |
| A-levels or vocational | 31.7% |  | 32.4% |  | 33.6% |  |
| Up to GCSE | 32.2% |  | 31.2% |  | 35.5% |  |
| *Relationship breakdowns, loss, and conflict* |  |  |  |  |  |  |
| Separation or divorce | 3.8% |  | 8.0% |  | 9.5% |  |
| Family problem | 16.4% |  | 29.1% |  | 31.8% |  |
| Other relationship breakdown | 11.3% |  | 21.1% |  | 28.1% |  |
| Death of close relative or friend | 32.0% |  | 33.1% |  | 37.8% |  |
| Mental health condition (ref none) | 15.5% |  | 38.7% |  | 58.8% |  |
| *Coping styles* |  |  |  |  |  |  |
| Problem-focused coping (range 0-12) | 4.38 | 0.03 | 4.75 | 0.07 | 4.61 | 0.13 |
| Emotion-focused coping (range 0-24) | 9.12 | 0.04 | 8.93 | 0.09 | 8.49 | 0.19 |
| Socially supported coping (range 0-18) | 4.40 | 0.04 | 5.37 | 0.08 | 5.54 | 0.17 |
| Avoidant coping (range 0-18) | 1.98 | 0.03 | 3.76 | 0.07 | 4.75 | 0.16 |

Note. Data were weighted to the proportions of gender, age, ethnicity, country, and education obtained from the Office for National Statistics. GCSE refers to General Certificate of Secondary Education.

Women, people from ethnic minority groups, and young adults were more likely to have thought about or engaged in self-harm at least once over the study period. Death of a close relative or friend was the most common relationship breakdown, loss, or conflict variable in the total sample and in people with self-harm thoughts or behaviours. People who had thought about or engaged in self-harm were more likely than the total sample to have been separated/divorced, had a family problem, or experienced another relationship breakdown in the past year. People with self-harm behaviours were slightly more likely to have experienced the death of a close relative or friend (37.8% vs total sample: 32.0% vs people with self-harm thoughts: 33.1%). A clinically diagnosed mental health condition, the use more problem-focused, socially supported, and avoidant coping mechanisms, and fewer emotion-focused coping strategies were also more common in participants with self-harm thoughts and behaviours than the total sample.

### Associations between relationship breakdowns, loss, and conflict with outcomes

Results from Poisson regressions predicting the frequency of self-harm thoughts and behaviours over the past year from relationship breakdowns, loss, and conflict are presented in Table 2. In fully adjusted models which included all four predictor variables as well as covariates, having had a family problem in the past year was the strongest predictor of more frequent self-harm thoughts (incidence rate ratio [IRR]: 1.77; 95% confidence interval [CI]: 1.71 to 1.83), whilst having experienced an ‘other’ relationship breakdown was the largest predictor of self-harm behaviours frequency (IRR: 1.96; 95% CI: 1.82 to 2.12). Associations were smallest for death of a close relative or friend.

**Table 2.** Associations between relationship breakdown variables and self-harm thoughts and behaviours frequency using Poisson regressions (N = 21,581), weighted

|  | Self-harm thoughts frequency |  |  |  |  |  | Self-harm behaviours frequency |  |  |  |  |  |
| --- | --- | --- | --- | --- | --- | --- | --- | --- | --- | --- | --- | --- |
|  | Unadjusted IRR | 95% CI |  | Adjusted IRR | 95% CI |  | Unadjusted IRR | 95% CI |  | Adjusted IRR | 95% CI |  |
| Separation or divorce | **1.57** | 1.48 | 1.65 | **1.31** | 1.24 | 1.38 | **2.36** | 2.13 | 2.61 | **1.82** | 1.64 | 2.01 |
| Family problem | **1.99** | 1.92 | 2.06 | **1.77** | 1.71 | 1.83 | **1.93** | 1.79 | 2.08 | **1.66** | 1.54 | 1.79 |
| Other relationship breakdown | **1.91** | 1.84 | 1.98 | **1.52** | 1.47 | 1.58 | **2.80** | 2.59 | 3.01 | **1.96** | 1.82 | 2.12 |
| Death of close relative or friend | 1.00 | 0.97 | 1.03 | **1.04** | 1.01 | 1.08 | **1.39** | 1.30 | 1.49 | **1.48** | 1.38 | 1.58 |

Note: Adjusted regression models included demographic variables and data were weighted to the proportions of gender, age, ethnicity, country, and education obtained from the Office for National Statistics.

### Moderation of associations between separation or divorce with outcomes

Having experienced a separation or divorce in the past year remained predictive of more frequent self-harm thoughts and behaviours over the study period (self-harm thoughts: IRR range: 1.77 to 1.82; self-harm behaviours: IRR range: 2.36 to 2.82) in models with interaction terms (Table 3). The use of more avoidant and socially supported coping strategies increased risk for both outcomes. Emotion-focused coping decreased the frequency of self-harm thoughts (IRR: 0.85; 95% CI: 0.81 to 0.90) and behaviours (IRR: 0.85; 95% CI: 0.73 to 0.98), but there was no evidence for an interaction between this coping strategy with separation or divorce. There was, however, evidence that greater use of avoidant coping strategies scores attenuated the association between separation or divorce and the frequency of self-harm thoughts, whilst problem-focused coping attenuated the impact of this exposure variable on self-harm behaviours.

**Table 3.** Interactions between separation or divorce and moderators predicting the frequency of self-harm thoughts and behaviours over the study period using Poisson regressions (N = 21,581), weighted

|  | Self-harm thoughts frequency |  |  | Self-harm behaviours frequency |  |  |
| --- | --- | --- | --- | --- | --- | --- |
|  | IRR | 95% CI |  | IRR | 95% CI |  |
| Separation or divorce | **1.77** | 1.47 | 2.14 | **2.56** | 1.60 | 4.11 |
| Problem-focused coping | **1.03** | 0.98 | 1.09 | 1.02 | 0.90 | 1.16 |
| Interaction with separation/divorce | 0.90 | 0.75 | 1.08 | **0.62** | 0.40 | 0.95 |
| Separation or divorce | **1.82** | 1.51 | 2.20 | **2.82** | 1.67 | 4.75 |
| Emotion-focused coping | **0.85** | 0.81 | 0.90 | **0.85** | 0.73 | 0.98 |
| Interaction with separation/divorce | 1.11 | 0.89 | 1.38 | 1.20 | 0.68 | 2.13 |
| Separation or divorce | **1.78** | 1.47 | 2.15 | **2.75** | 1.67 | 4.54 |
| Socially supported coping | **1.13** | 1.07 | 1.19 | **1.13** | 1.01 | 1.27 |
| Interaction with separation/divorce | 0.91 | 0.78 | 1.06 | 0.90 | 0.69 | 1.16 |
| Separation or divorce | **1.82** | 1.44 | 2.29 | **2.36** | 1.07 | 5.21 |
| Avoidant coping | **1.54** | 1.49 | 1.59 | **1.63** | 1.53 | 1.74 |
| Interaction with separation/divorce | **0.83** | 0.76 | 0.91 | 0.92 | 0.75 | 1.14 |

Note. Poisson regression models included demographic variables and data were weighted to the proportions of gender, age, ethnicity, country, and education obtained from the Office for National Statistics.

### Moderation of associations between a family problem with outcomes

Having had a family problem in the past year continued to predict the frequency of both outcomes once moderators and their interactions were included in models (self-harm thoughts IRR range: 1.92 to 2.03; self-harm behaviours IRR range: 1.93 to 2.17) (Table 4). The use of avoidant and socially supported coping strategies also increased the likelihood of both outcomes, whilst emotion-focused coping decreased the frequency of self-harm thoughts (IRR: 0.84; 95% CI: 0.79 to 0.89). However, as with separation or divorce, avoidant coping strategies attenuated the association between having had a family problem and self-harm thoughts (avoidant coping strategies: IRR: 0.85; 95% CI: 0.81 to 0.90). The only interaction found between having had a family problem and self-harm behaviours frequency was with problem-focused coping (IRR: 0.74; 95% CI: 0.57 to 0.95).

**Table 4.**
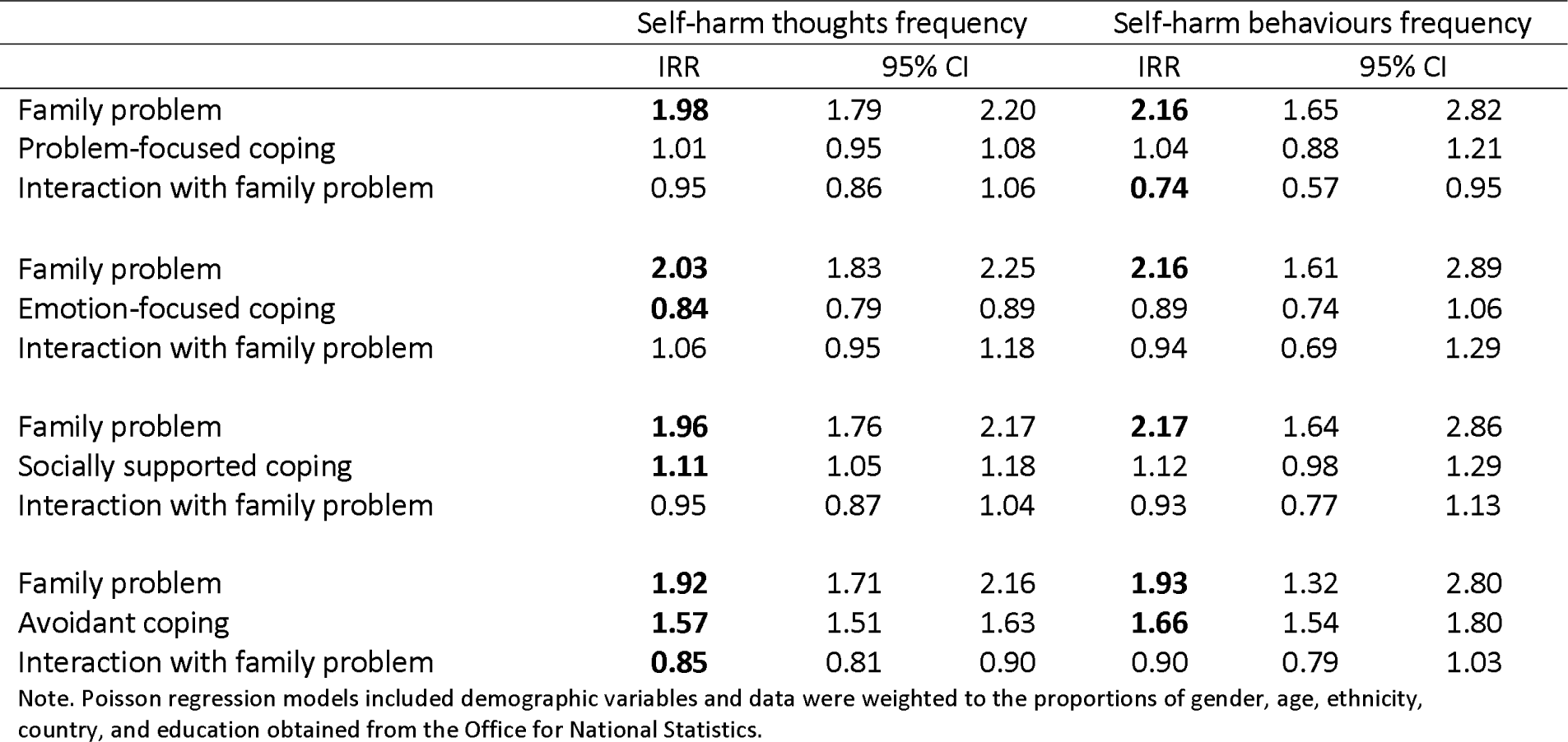
Interactions between a family problem and moderators predicting the frequency of self-harm thoughts and behaviours using Poisson regressions (N = 21,581), weighted

|  | Self-harm thoughts frequency |  |  | Self-harm behaviours frequency |  |  |
| --- | --- | --- | --- | --- | --- | --- |
|  | IRR | 95% CI |  | IRR | 95% CI |  |
| Family problem | **1.98** | 1.79 | 2.20 | **2.16** | 1.65 | 2.82 |
| Problem-focused coping | 1.01 | 0.95 | 1.08 | 1.04 | 0.88 | 1.21 |
| Interaction with family problem | 0.95 | 0.86 | 1.06 | **0.74** | 0.57 | 0.95 |
| Family problem | **2.03** | 1.83 | 2.25 | **2.16** | 1.61 | 2.89 |
| Emotion-focused coping | **0.84** | 0.79 | 0.89 | 0.89 | 0.74 | 1.06 |
| Interaction with family problem | 1.06 | 0.95 | 1.18 | 0.94 | 0.69 | 1.29 |
| Family problem | **1.96** | 1.76 | 2.17 | **2.17** | 1.64 | 2.86 |
| Socially supported coping | **1.11** | 1.05 | 1.18 | 1.12 | 0.98 | 1.29 |
| Interaction with family problem | 0.95 | 0.87 | 1.04 | 0.93 | 0.77 | 1.13 |
| Family problem | **1.92** | 1.71 | 2.16 | **1.93** | 1.32 | 2.80 |
| Avoidant coping | **1.57** | 1.51 | 1.63 | **1.66** | 1.54 | 1.80 |
| Interaction with family problem | **0.85** | 0.81 | 0.90 | 0.90 | 0.79 | 1.03 |

Note. Poisson regression models included demographic variables and data were weighted to the proportions of gender, age, ethnicity, country, and education obtained from the Office for National Statistics.

### Moderation of associations between another relationship breakdown with outcomes

The breakdown of an ‘other’ relationship increased the frequency of both outcomes (self-harm thoughts IRR range: 1.72 to 1.87; self-harm behaviours IRR range: 1.84 to 2.66), as did avoidant and socially supported coping, whilst emotion-focused coping reduced the likelihood of both outcomes (Table 5). As with separation/divorce and having had a family problem, the association between having an ‘other’ relationship breakdown with self-harm thoughts was attenuated by the use of more avoidant and socially supported coping strategies (avoidant coping: IRR: 0.86; 95% CI: 0.82 to 0.92). Socially supported coping attenuated the impact of having had an ‘other’ relationship breakdown on both outcomes. Emotion-focused coping exacerbated the association of having had an ‘other’ relationship breakdown with self-harm thoughts: IRR: 1.18; 95% CI: 1.05; 95% CI: 1.32).

**Table 5.** Interactions between having an ‘other’ relationship breakdown and moderators predicting the frequency of self-harm thoughts and behaviours using Poisson regressions (N = 21,581), weighted

|  | Self-harm thoughts frequency |  |  | Self-harm behaviours frequency |  |  |
| --- | --- | --- | --- | --- | --- | --- |
|  | IRR | 95% CI |  | IRR | 95% CI |  |
| Other relationship breakdown | **1.79** | 1.60 | 2.00 | **2.54** | 1.97 | 3.29 |
| Problem-focused coping | 1.00 | 0.94 | 1.06 | 0.91 | 0.80 | 1.05 |
| Interaction with other relationship breakdown | 1.02 | 0.92 | 1.14 | 1.05 | 0.81 | 1.35 |
| Other relationship breakdown | **1.87** | 1.68 | 2.09 | **2.66** | 2.04 | 3.46 |
| Emotion-focused coping | **0.82** | 0.78 | 0.87 | **0.79** | 0.69 | 0.91 |
| Interaction with other relationship breakdown | **1.18** | 1.05 | 1.32 | 1.30 | 0.93 | 1.81 |
| Other relationship breakdown | **1.80** | 1.61 | 2.01 | **2.57** | 1.99 | 3.33 |
| Socially supported coping | **1.13** | 1.06 | 1.20 | **1.19** | 1.03 | 1.37 |
| Interaction with other relationship breakdown | **0.90** | 0.82 | 0.99 | **0.77** | 0.63 | 0.95 |
| Other relationship breakdown | **1.72** | 1.51 | 1.95 | **1.84** | 1.36 | 2.49 |
| Avoidant coping | **1.55** | 1.50 | 1.61 | **1.58** | 1.46 | 1.71 |
| Interaction with other relationship breakdown | **0.86** | 0.82 | 0.92 | 1.00 | 0.89 | 1.13 |

Note. Poisson regression models included demographic variables and data were weighted to the proportions of gender, age, ethnicity, country, and education obtained from the Office for National Statistics.

### Moderation of associations between death of a close relative or friend with outcomes

Death of a close relative or friend increased the frequency of self-harm thoughts (IRR range: 1.12 to 1.15), and behaviours in most models with interaction terms (IRR range: 1.47 to 1.73) (Table 6). The use of more socially supported and avoidant coping strategies also increased the frequency of both outcomes, whilst emotion-focused coping reduced this risk. There was no evidence for interactions between the moderators and having experienced the death of a close relative or friend.

**Table 6.** Interactions between death of close relative or friend and moderators predicting the frequency of self-harm thoughts and behaviours using Poisson regressions (N = 21,581), weighted

|  | Self-harm thoughts frequency |  |  | Self-harm behaviours frequency |  |  |
| --- | --- | --- | --- | --- | --- | --- |
|  | IRR | 95% CI |  | IRR | 95% CI |  |
| Death of close relative or friend | **1.12** | 1.00 | 1.25 | **1.67** | 1.29 | 2.15 |
| Problem-focused coping | 1.01 | 0.95 | 1.07 | 0.93 | 0.81 | 1.06 |
| Interaction with death of close relative or friend | 1.04 | 0.94 | 1.16 | 1.08 | 0.84 | 1.40 |
| Death of close relative or friend | **1.15** | 1.03 | 1.29 | **1.73** | 1.33 | 2.26 |
| Emotion-focused coping | **0.85** | 0.80 | 0.90 | **0.81** | 0.69 | 0.96 |
| Interaction with death of close relative or friend | 1.05 | 0.94 | 1.17 | 1.15 | 0.87 | 1.53 |
| Death of close relative or friend | 1.10 | 0.99 | 1.23 | **1.63** | 1.26 | 2.10 |
| Socially supported coping | **1.11** | 1.05 | 1.18 | **1.12** | 0.98 | 1.28 |
| Interaction with death of close relative or friend | 1.03 | 0.93 | 1.13 | 0.99 | 0.81 | 1.21 |
| Death of close relative or friend | 1.04 | 0.92 | 1.18 | **1.47** | 1.07 | 2.02 |
| Avoidant coping | **1.52** | 1.46 | 1.57 | **1.62** | 1.50 | 1.74 |
| Interaction with death of close relative or friend | 0.99 | 0.94 | 1.05 | 1.01 | 0.89 | 1.13 |

Note. Poisson regression models included demographic variables and data were weighted to the proportions of gender, age, ethnicity, country, and education obtained from the Office for National Statistics.

### Sensitivity analyses

In models with predictor variables comparing those to whom the event did not happen, or who were not upset too much or at all vs those who were moderately or very much upset, substantive results were the same as in the main analyses (Supplemental Tables S4-S8). When the ‘venting’ items were removed from the socially supported coping scale, this coping strategy reduced the likelihood of outcomes in most models, rather than increasing risk as they did in the main analyses when the venting items were included in this scale (Supplemental Table S9). Socially supported coping also attenuated the impact of several predictor variables (e.g., separation or divorce, family problem) on outcomes, but more so for self-harm behaviours.

## Discussion

Using longitudinal data from a large sample of UK adults, we examined whether relationship breakdowns, loss, and conflict increased the frequency of self-harm thoughts and behaviours over the course of the second year of the COVID-19 pandemic. Then, we investigated whether different coping styles, suggested by theoretical models of self-harm risk, interacted with these stressors in relation to these outcomes. All four relationship variables generally increased the risk for more frequent self-harm thoughts and behaviours, as did the use of more avoidant coping strategies (e.g., substance use and denial). Emotion-focused coping (e.g., positive reframing, acceptance, humour) reduced the risk for more frequent self-harm thoughts and behaviours. However, results from interactions between relationship breakdowns, loss, and conflict with these factors provided mixed support for theoretical models; avoidant coping attenuated the impact of predictor variables on self-harm thoughts (but not behaviours).

Amongst the four variables representing relationship dissolution, loss, and conflict increased the frequency of outcomes, associations were stronger for family problems, separation or divorce, and the breakdown of an ‘other’ relationship compared to those for death of a close relative or friend. This is consistent with research showing romantic relationship difficulties and family problems as the most frequent stressors precipitating self-harm.^6, 7^ Stress as a result of the COVID-19 pandemic may have had a ‘spillover effect’ into interpersonal relationships, placing pressure on dyadic and individual coping resources.^16, 17^ The pandemic has also likely placed additional strains on family members and romantic partners to meet social needs that were previously met elsewhere (e.g., in the workplace) when restrictions on face-to-face contact were not in place. Although the use of more avoidant coping strategies increased the risk for the frequency of self-harm thoughts and behaviours, they attenuated risk for self-harm thoughts in interaction with many of the exposure variables, but not for self-harm behaviours. It could be that the use of these coping strategies captured in the scale used, which were relinquishing efforts to deal with the situation, engaging in denial, or using substances to cope, were helpful in the short term for reducing the psychological stress associated with these events and redirecting thoughts away from those stressors. But it is notable that there was no protective association with self-harm behaviours, reinforcing previous research suggesting that such coping strategies are not adaptive in the long-term.

Another key finding was that emotion-focused coping exacerbated the impact of having had a breakdown of an ‘other’ relationship on self-harm thoughts. This was not entirely unexpected, as emotion-oriented coping has been found to be associated with self-harm thoughts and behaviours in adolescents.^26^ The emotion-focused coping scale used in the current study involved reframing the event more positively, using humour to deal with the situation, and using religious or spiritual beliefs or practices to cope. However, in the context of the COVID-19 pandemic, such strategies may have been insufficient for coping with a stressor as significant as a relationship breakdown, which could have involved having to make plans to relocate or change one’s living circumstances, for which opportunities were limited due to restrictions on household mixing and moving house.

Additionally, we found that when the socially supported coping scale included venting (e.g., saying things to let one’s unpleasant feelings escape), this way of coping increased risk for both outcomes. This is not entirely surprising, given prior work showing that venting negatively impacts mental health.^12, 22^ However, when the two venting items were removed, this way of coping reduced the frequency of both outcomes, and attenuated the impact of several predictors (e.g., divorce or separation, family problem) on outcomes, as predicted by previous literature. ^12, 22^ The remaining four items comprising the socially supported coping scale reflected emotional (e.g., getting comfort and understanding from someone) and instrumental support (e.g., getting help and advice from other people). Inadequate social support in the context of psychological distress is a key aspect of theoretical models of self-harm risk.^8, 9^ In the UK Biobank study, a lack of emotional support explained longitudinal associations between living alone and later death by suicide in men (but not women).^27^ Conversely, higher emotional support is associated with reduced likelihood of self-harm thoughts.^28^

This study has several strengths, including the use of a large well-stratified sample and repeated measures of the outcome variables. However, there are also several limitations. First, due to data limitations, we did not include measures of other factors thought to form the diathesis for suicide and self-harm risk, such as impulsivity and proneness to aggression.^10, 29^ Second, although the wording of our self-harm behaviours question was developed to be similar to our self-harm thoughts question, the way the question was phrased (“How often have you been bothered by…”) may have led to underreporting of self-harm behaviours. This is because first-hand accounts of reasons for self-harm suggest the behaviour is often perceived positively and used as a coping mechanism by individuals who self-harm.^30^ The same can be said of thoughts of self-harm or suicide, which can, like self-harm behaviours, provide an individual experiencing distress with a sense of soothing and self-regulation.^31^ Although well-stratified across major demographic groups and heterogenous, the sample was not representative of the UK general population. We may have therefore failed to capture individuals who were experiencing the most extreme distress. Further, although this study included regular monthly follow-ups, there is still a possibility that some who were excluded were engaging in more severe self-harm thoughts and behaviours and were therefore unable to participate. However, the comparison of excluded vs included participants did not indicate that the former group had experienced more relationship breakdowns or self-harm thoughts and behaviours. Further research is also needed to unpack the heterogeneity within the ‘other’ relationship breakdown category, as question wording prevented us from considering this. Finally, whilst this study gives particular insight into circumstances surrounding relationship breakdowns and self-harm during the COVID-19 pandemic, replication in non-COVID times is needed.

## Conclusions

Our findings demonstrate the importance of interpersonal loss and conflict for more frequent self-harm thoughts and behaviours during the second year of the COVID-19 pandemic. Avoidant coping strategies increased the risk for self-harm thoughts and behaviours. However, we found little evidence from statistical interactions to support predictions from theoretical models^8, 9^ that these long-standing factors increase the likelihood of thinking about and engaging in self-harm when these stressful life events occur. Most people who experience breakdowns or conflict in relationships do not think about or engage in self-harm, so ongoing research to understand the factors which attenuate or exacerbate this risk is a research priority.

## Data Availability

Data produced will be publicly available at the end of 2022.

## Declarations

### Ethics approval and consent to participate

Ethical approval for the COVID-19 Social Study was granted by the UCL Ethics Committee [approval number 12467/005]. The study was performed in accordance with the Declaration of Helsinki. Participants were not compensated for participation. All participants provided fully informed consent and the study is GDPR compliant.

### Consent for publication

Not applicable.

### Availability of data and materials

The dataset analysed for the current study is not yet publicly available due to funder restrictions. However, the UCL COVID-19 Social Study documentation and codebook are available for download at https://osf.io/jm8ra/. Statistical code is available upon request from Elise Paul.

### Competing interests

The authors declare that they have no competing interests.

### Funding

The Nuffield Foundation [WEL/FR-000022583], the MARCH Mental Health Network funded by the Cross-Disciplinary Mental Health Network Plus initiative supported by UK Research and Innovation [ES/S002588/1], and the Wellcome Trust [221400/Z/20/Z and 205407/Z/16/Z].

### Author contributions

EP conceptualised and designed the current study. DF acquired funding, provided oversight on the methodology, administered the project, provided software and other resources, and supervised the project. Data were curated, validated, and formally analysed by EP. EP created visualisations, wrote the original manuscript draft with input from all authors. Both authors reviewed and edited the manuscript.

## Acknowledgements

The researchers are grateful for the support of a number of organisations with their recruitment efforts including: the UKRI Mental Health Networks, Find Out Now, UCL BioResource, SEO Works, FieldworkHub, and Optimal Workshop.

## Supplemental Materials

**Table S1.**
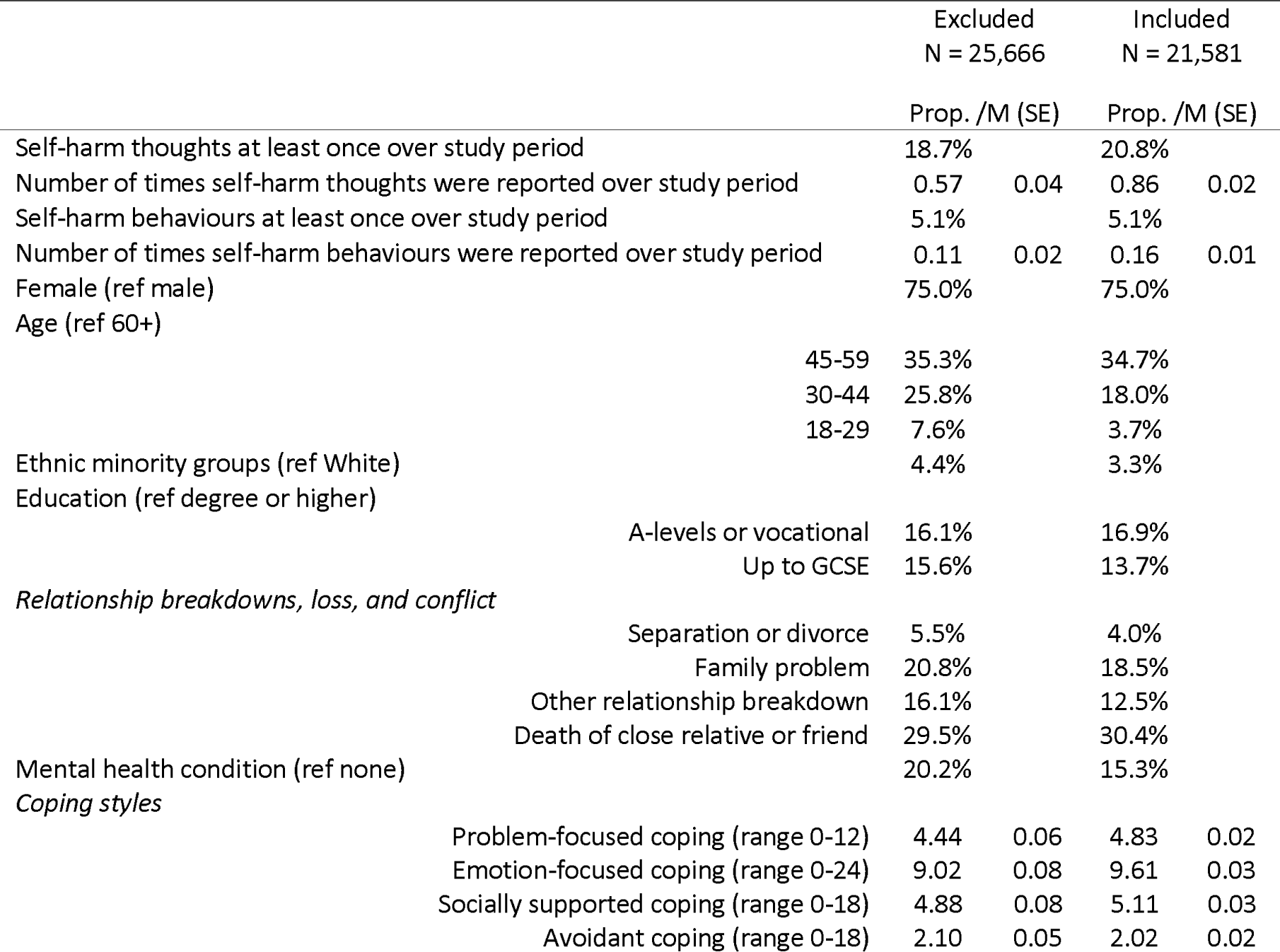
Characteristics of excluded and included participants, unweighted

**Table S2.**
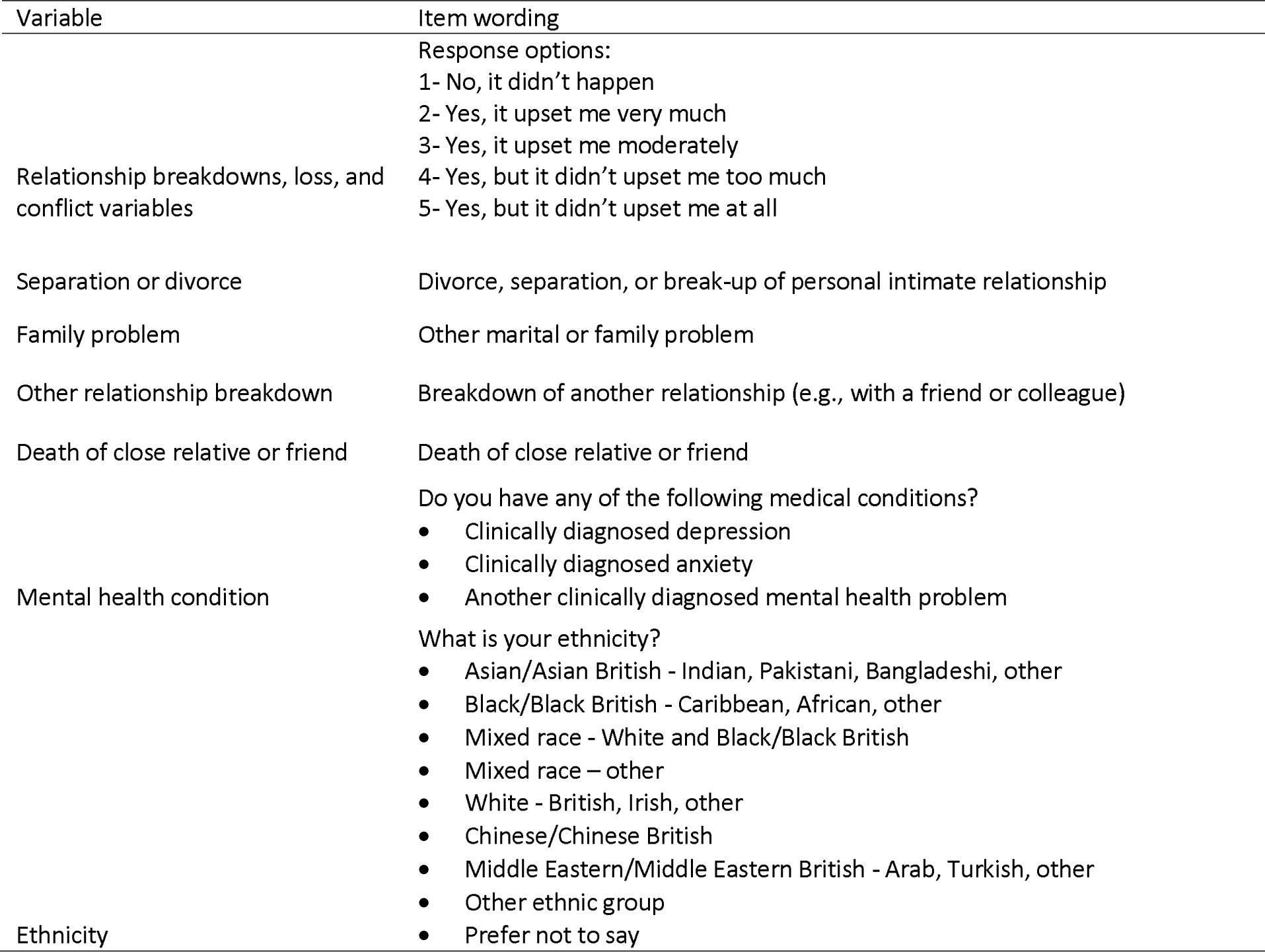
Study developed variables

**Table S3.** Weighted and unweighted sample characteristics (N = 21,581)

|  | Unweighted |  | Weighted |  |
| --- | --- | --- | --- | --- |
|  | Prop. | /M (SE) | Prop. | /M (SE) |
| Self-harm thoughts at least once over study period | 20.8% |  | 21.1% |  |
| Number of times self-harm thoughts were reported over study period | 0.86 | 0.02 | 0.94 | 0.03 |
| Self-harm behaviours at least once over study period | 5.1% |  | 5.6% |  |
| Number of times self-harm behaviours were reported over study period | 0.16 | 0.01 | 0.19 | 0.01 |
| Female (ref male) | 75.0% |  | 50.8% |  |
| Age (ref 60+) |  |  |  |  |
| 45-59 | 34.7% |  | 29.1% |  |
| 30-44 | 18.0% |  | 14.4% |  |
| 18-29 | 3.7% |  | 5.8% |  |
| Ethnic minority groups (ref White) | 3.3% |  | 6.5% |  |
| Education (ref degree or higher) |  |  |  |  |
| A-levels or vocational | 16.9% |  | 31.7% |  |
| Up to GCSE | 13.7% |  | 32.2% |  |
| *Relationship breakdowns, loss, and conflict* |  |  |  |  |
| Separation or divorce | 4.0% |  | 3.8% |  |
| Family problem | 18.5% |  | 16.4% |  |
| Other relationship breakdown | 12.5% |  | 11.3% |  |
| Death of close relative or friend | 30.4% |  | 32.0% |  |
| Mental health condition (ref none) | 15.3% |  | 15.5% |  |
| *Coping styles* |  |  |  |  |
| Problem-focused coping (range 0-12) | 4.83 | 0.02 | 4.38 | 0.03 |
| Emotion-focused coping (range 0-24) | 9.61 | 0.03 | 9.12 | 0.04 |
| Socially supported coping (range 0-18) | 5.11 | 0.03 | 4.40 | 0.04 |
| Avoidant coping (range 0-18) | 2.02 | 0.02 | 1.98 | 0.03 |

Note. Data in the weighted sample were weighted to the proportions of gender, age, ethnicity, country, and education obtained from the Office for National Statistics. GCSE refers to General Certificate of Secondary Education.

**Table S4.**
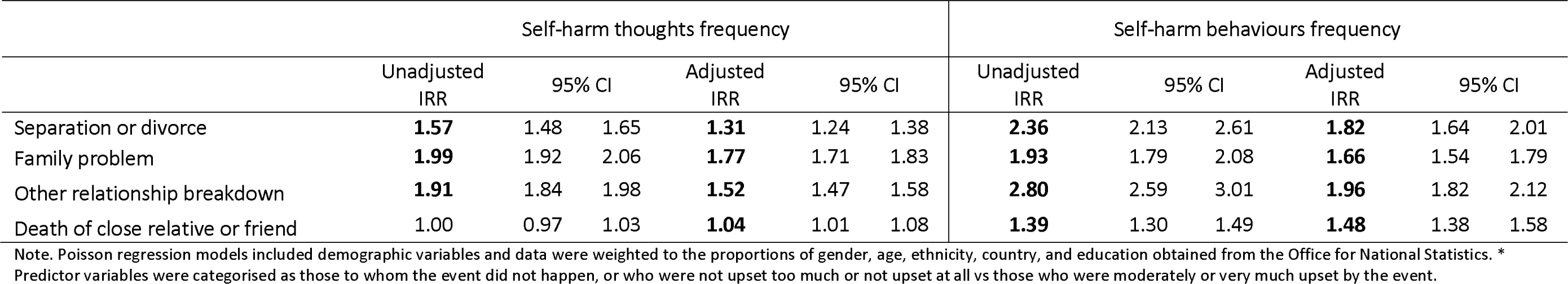
Sensitivity analysis: Associations between relationship breakdowns, loss, and conflict* with self-harm thoughts and behaviours frequency using Poisson regressions (N = 21,581), weighted

**Table S5.** Sensitivity analysis: Interactions between separation or divorce* and moderators predicting the frequency of self-harm thoughts and behaviours over the study period using Poisson regressions (N = 21,581), weighted

|  | Self-harm thoughts frequency |  |  | Self-harm behaviours frequency |  |  |
| --- | --- | --- | --- | --- | --- | --- |
|  | IRR | 95% CI |  | IRR | 95% CI |  |
| Separation or divorce | **1.89** | 1.57 | 2.27 | **2.38** | 1.57 | 3.60 |
| Problem-focused coping | 1.03 | 0.98 | 1.09 | 1.00 | 0.88 | 1.14 |
| Interaction with separation/divorce | 0.89 | 0.73 | 1.07 | 0.67 | 0.40 | 1.10 |
| Separation or divorce | **1.93** | 1.61 | 2.32 | **2.51** | 1.62 | 3.90 |
| Emotion-focused coping | **0.85** | 0.81 | 0.90 | **0.84** | 0.73 | 0.97 |
| Interaction with separation/divorce | 1.12 | 0.89 | 1.42 | 1.35 | 0.67 | 2.75 |
| Separation or divorce | **1.92** | 1.59 | 2.31 | **2.52** | 1.63 | 3.89 |
| Socially supported coping | **1.13** | 1.07 | 1.19 | **1.14** | 1.01 | 1.28 |
| Interaction with separation/divorce | 0.87 | 0.74 | 1.03 | 0.82 | 0.61 | 1.10 |
| Separation or divorce | **1.90** | 1.53 | 2.35 | 1.67 | 0.97 | 2.90 |
| Avoidant coping | **1.54** | 1.49 | 1.58 | **1.61** | 1.51 | 1.72 |
| Interaction with separation/divorce | **0.83** | 0.75 | 0.91 | 1.00 | 0.84 | 1.20 |

Note. Poisson regression models included demographic variables and data were weighted to the proportions of gender, age, ethnicity, country, and education obtained from the Office for National Statistics. \* Predictor variables were categorised as those to whom the event did not happen, or who were not upset too much or not upset at all vs those who were moderately or very much upset by the event.

**Table S6.** Sensitivity analysis: Interactions between a family problem* and moderators predicting the frequency of self-harm thoughts and behaviours using Poisson regressions (N = 21,581), weighted

|  | Self-harm thoughts frequency |  |  | Self-harm behaviours frequency |  |  |
| --- | --- | --- | --- | --- | --- | --- |
|  | IRR | 95% CI |  | IRR | 95% CI |  |
| Family problem | **1.97** | 1.77 | 2.18 | **2.04** | 1.59 | 2.61 |
| Problem-focused coping | 1.03 | 0.97 | 1.09 | 1.02 | 0.88 | 1.19 |
| Interaction with family problem | 0.92 | 0.83 | 1.02 | **0.76** | 0.58 | 0.99 |
| Family problem | **1.99** | 1.79 | 2.22 | **2.01** | 1.52 | 2.67 |
| Emotion-focused coping | **0.85** | 0.80 | 0.90 | 0.89 | 0.75 | 1.05 |
| Interaction with family problem | 1.04 | 0.93 | 1.16 | 0.94 | 0.67 | 1.32 |
| Family problem | **1.96** | 1.76 | 2.18 | **2.08** | 1.61 | 2.68 |
| Socially supported coping | **1.13** | 1.06 | 1.19 | **1.15** | 1.01 | 1.32 |
| Interaction with family problem | 0.92 | 0.84 | 1.01 | 0.86 | 0.71 | 1.06 |
| Family problem | **1.98** | 1.77 | 2.22 | **1.69** | 1.25 | 2.28 |
| Avoidant coping | **1.57** | 1.52 | 1.63 | **1.63** | 1.50 | 1.76 |
| Interaction with family problem | **0.83** | 0.79 | 0.88 | 0.96 | 0.85 | 1.08 |

Note. Poisson regression models included demographic variables and data were weighted to the proportions of gender, age, ethnicity, country, and education obtained from the Office for National Statistics. \* Predictor variables were categorised as those to whom the event did not happen, or who were not upset too much or not upset at all vs those who were moderately or very much upset by the event.

**Table S7.**
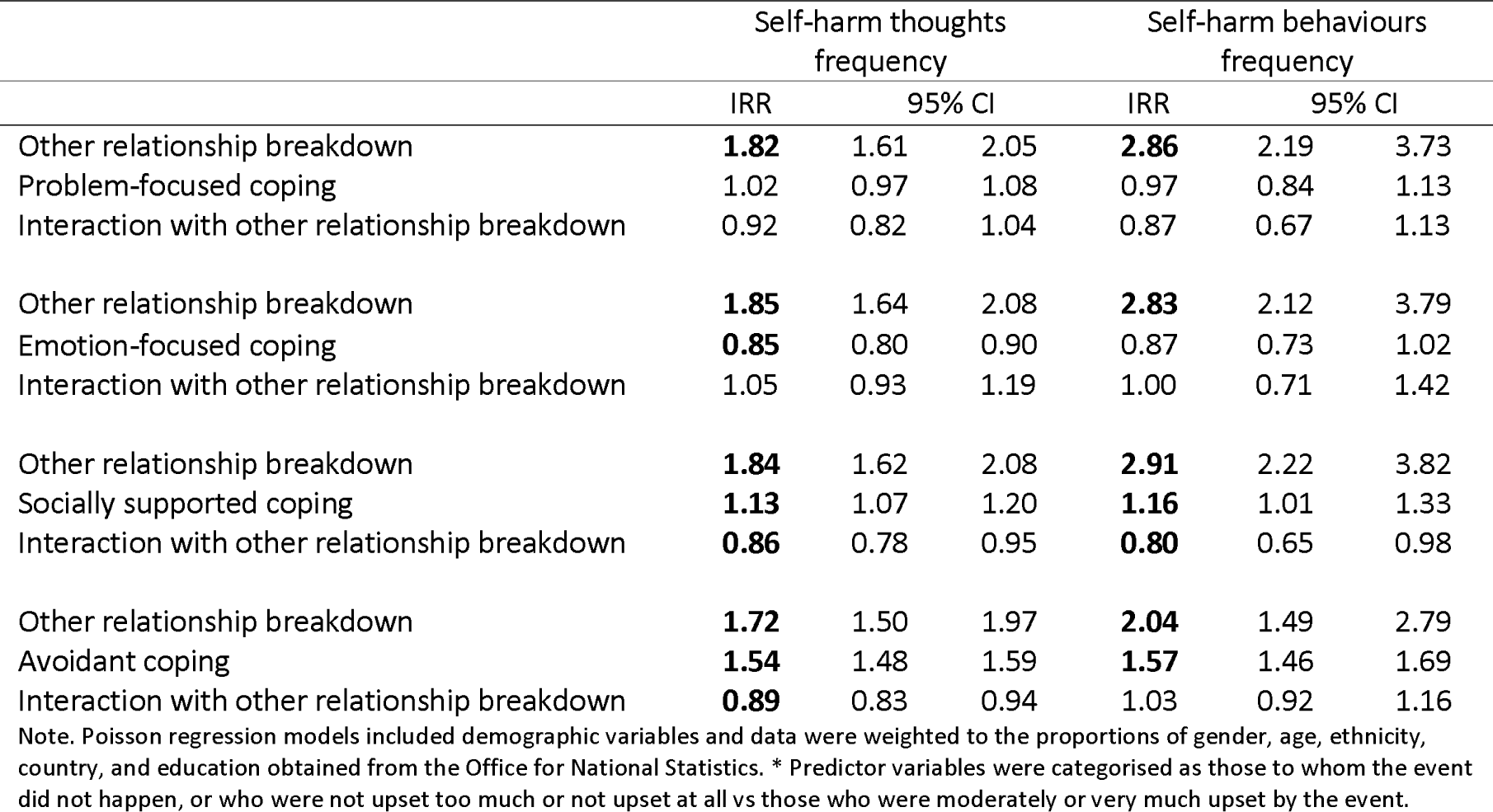
Interactions between having an ‘other’ relationship breakdown* and moderators predicting the frequency of self-harm thoughts and behaviours using Poisson regressions (N = 21,581), weighted

**Table S8.**
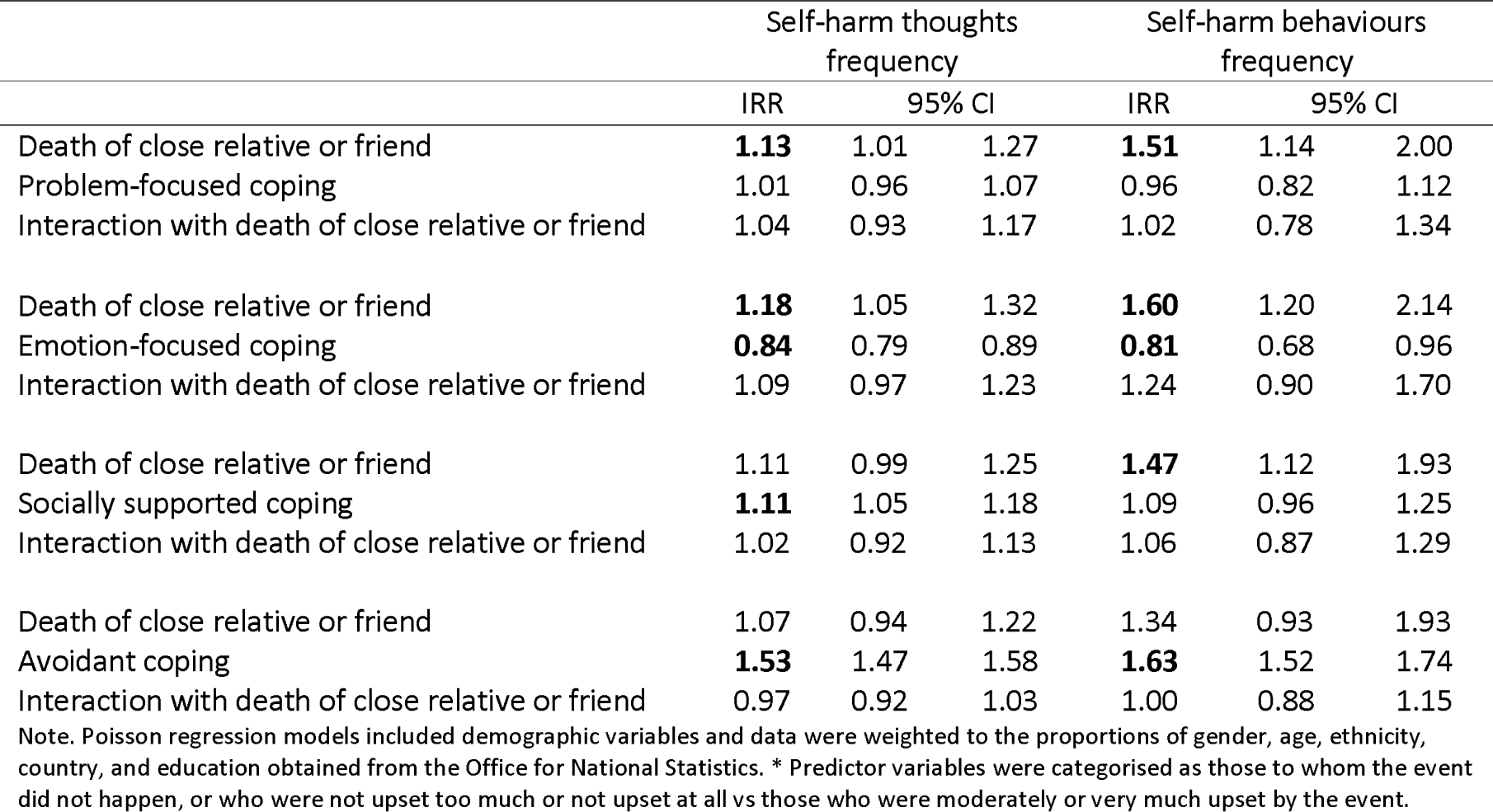
Interactions between death of close relative or friend and moderators predicting the frequency of self-harm thoughts and behaviours using Poisson regressions (N = 21,581), weighted

**Table S9.** Interactions between death of close relative or friend and socially supported coping (without venting) predicting the frequency of self-harm thoughts and behaviours using Poisson regressions (N = 21,581), weighted

|  | Self-harm thoughts frequency |  |  | Self-harm behaviours frequency |  |  |
| --- | --- | --- | --- | --- | --- | --- |
|  | IRR | 95% CI |  | IRR | 95% CI |  |
| Divorce or separation | **1.78** | 1.69 | 1.88 | **2.69** | 2.44 | 2.96 |
| Socially supported coping | **0.96** | 0.94 | 0.97 | **0.95** | 0.92 | 0.99 |
| Interaction with divorce or separation | **0.89** | 0.84 | 0.94 | **0.74** | 0.67 | 0.82 |
| Family problem | **1.99** | 1.93 | 2.06 | **2.17** | 2.02 | 2.33 |
| Socially supported coping | **0.94** | 0.93 | 0.96 | 0.95 | 0.91 | 1.00 |
| Interaction with family problem | 0.98 | 0.95 | 1.01 | **0.88** | 0.82 | 0.95 |
| Other relationship breakdown | **1.82** | 1.75 | 1.88 | **2.47** | 2.30 | 2.66 |
| Socially supported coping | **0.94** | 0.92 | 0.96 | 0.99 | 0.95 | 1.04 |
| Interaction with other relationship breakdown | 0.99 | 0.96 | 1.03 | **0.77** | 0.71 | 0.82 |
| Death of close relative or friend | **1.12** | 1.08 | 1.16 | **1.64** | 1.53 | 1.76 |
| Socially supported coping | **0.96** | 0.94 | 0.98 | **0.95** | 0.90 | 0.99 |
| Interaction with death of close relative or friend | 0.97 | 0.94 | 1.00 | 0.94 | 0.87 | 1.00 |

Note. Poisson regression models included demographic variables and data were weighted to the proportions of gender, age, ethnicity, country, and education obtained from the Office for National Statistics.

## Notes

### Competing Interest Statement

The authors have declared no competing interest.

### Author Declarations

Ethical approval for the COVID-19 Social Study was granted by the University College London Ethics Committee [approval number 12467/005]. The study was performed in accordance with the Declaration of Helsinki.

